# Clinical features of cystic neutrophil granulomatous mastitis in 62 cases

**DOI:** 10.1101/2023.12.08.23299512

**Authors:** Mengjie Wang, Dongxiao Zhang, Na Fu, Min Liu, Hongkai Zhang, Shuo Feng, Yifei Zeng, Wenjie Zhao, Jianchun Cui, Khattak Mazher Mansoor

**Author notes:** Correspondence; Department of Galactophore, Beijing Hospital of Traditional Chinese Medicine, Capital Medical University, No. 23, Art Museum Back Street, Dongcheng District, Beijing, People’s Republic of China. Contributing authors: MW; NF; ML; HZ; SF; YZ; WJ; JC; KM.

## Abstract

**Background:** Cystic Neutrophilic Granulomatous Mastitis (CNGM) is a rare inflammatory condition affecting the breast. Despite its rarity, understanding its pathogenesis and clinical features is crucial for accurate diagnosis and effective management. This study delves into the nuanced aspects of CNGM, shedding light on its unique characteristics and potential underlying mechanisms. Methods: In this meticulous investigation, we meticulously examined and analyzed the biological data, clinical features, ultrasound imaging findings, and histopathological morphological information of 62 patients diagnosed with CNGM after thorough pathological examination. The study cohort was sourced from the Galactophore department of Beijing Hospital of Traditional Chinese Medicine, spanning the period from September 2019 to September 2022. Results: Sixty-two patients, with an average age of 33.30 years, were predominantly female. Among the 52 patients with detailed documentation of onset following the final delivery, various factors were identified, including hyperprolactinemia, pituitary tumors, psychiatric medication history, granulomatous mastitis history, breast trauma history, and a family history of breast cancer. The primary clinical manifestations were characterized by pain and palpable masses, accompanied by localized symptoms such as redness, ulceration, nipple discharge, and nipple retraction. Additionally, systemic symptoms, such as fever, headache, erythema nodosum, and cough, were observed. Ultrasound examinations revealed predominantly hypoechoic masses with heterogeneous echogenicity. Axillary lymphadenopathy, dilated ducts, and thickening of breast tissue were also noted in some cases. Histopathological analyses demonstrated lobular structural destruction, acute and chronic inflammatory cell infiltration, multinucleated giant cell reactions, granulomas, and cyst formation. Gram staining revealed detection rates of 41.94% (26/62) for gram-positive bacteria and 11.29% (7/62) for gram-negative bacteria. Conclusion: This study highlights the occurrence of Chronic Nonspecific Granulomatous Mastitis (CNGM) in women of childbearing age. Factors such as milk stasis, mammary duct secretion overcharge, exogenous trauma, hormonal influences, and bacterial colonization are implicated in the initiation and recurrence of CNGM. Notably, nipple retraction emerged not only as a clinical symptom but also as a potential risk factor for CNGM. The prevalence of multiple hypoechoic regions in CNGM surpassed that observed in breast cancer cases. The detection of gram-positive bacteria underscores the pivotal role of bacterial infections in the development of CNGM.

## Introduction

Cystic neutrophilic granulomatous mastitis (CNGM) is a class of benign breast diseases closely related to bacterial infection and has a unique histological morphology^[1]^. Paviour et al.(2002) first identified the histological features of CNGM in 24 women: a highly unique histological pattern - a purulent lipogranuloma consisting of a central lipid vacuole surrounded by neutrophils and an outer cuff of epithelioid histiocytes, mixed inflammatory infiltrates including Langerhans giant cells, lymphocytes^[2]^. Renshaw et al. defined this histological pattern as “cystic neutrophil granulomatous mastitis” in 2011^[3]^. In recent years, a series of studies on CNGM has given us a preliminary understanding of this disease. However, problems remain, such as unclear pathogenesis, preliminary diagnosis, and confusing treatment. We hope this study on the clinical characteristics of CNGM will help to understand this disease better.

CNGM was previously considered a subtype of GLM^[4-7]^. After further study, some scholars believe that CNGM is quite different from GLM in terms of Corynebacteria’s histological morphology and detection rate, so it is regarded as an independent disease^[8]^. But other scholars believe that CNGM is an infectious disease with histomorphological evolution. Signature lipid vacuoles have also been developed and may appear as a “fence” in the early stage^[9]^. This view is consistent with Oddo’s idea that there may be morphological patterns of evolution in bacteria-associated mastitis^[10]^. At present, the understanding of CNGM is not sufficient. Only about 300 cases of CNGM have been reported in the world. Most of the research is small sample size studies, and the universality of the results is not enough. According to the literature we retrieved, this study is currently the most extensive sample size study for the characteristic clinical analysis of CNGM.

This article aims to analyze the data of 62 CNGM cases and elaborate on and summarize their biological characteristics, clinical features, imaging manifestations, and histopathological morphology. The purpose is to have a deeper insight into CNGM and provide guidance for the early diagnosis and effective treatment of CNGM.

## Patients and methods

### Patients

In this study, we observed 62 patients diagnosed with CNGM according to the diagnostic criteria in “Cystic neutrophilic granulomatous mastitis: an update”^[1]^. All patients were from the Galactophore Department of Beijing Hospital of Traditional Chinese Medicine from September 2019 to September 2022. (shown in Fig. 1)

**Fig. 1.**
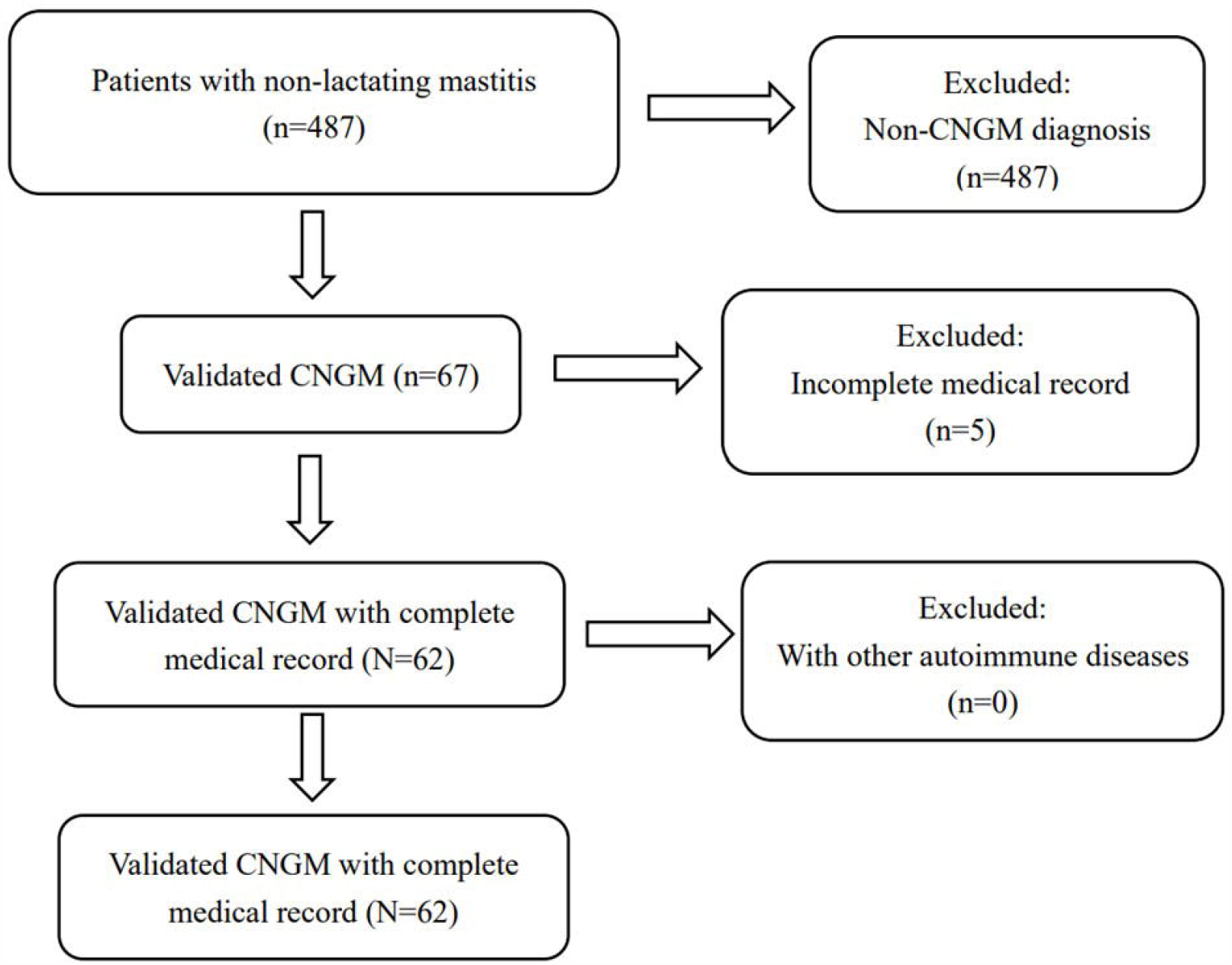
Flowchart for selecting patients.

Inclusion criteria: (1) The pathological diagnosis of the patients was CNGM; (2) People with complete clinical data.

Exclusion criteria: (1) Incomplete data: more than 30% of clinical data were missing; (2) People with other autoimmune diseases(such as systemic lupus erythematosus, and rheumatism).

### Research methods

Our team completed a retrospective statistical review of clinical data from 62 cases of CNGM patients between May 2023 and June 2023. The data of 62 patients with CNGM were summarized and cross-checked separately by two statistical analysts with the help of EXCEL in this study. The statistical processing was carried out through SPSS 25.0. Quantitative data were described as the means ± standard deviations, and qualitative data were expressed as frequencies and percentages. The biological data, clinical features, imaging manifestations, and histopathological morphology were analyzed in detail.

### Ethics Statement

This study was approved by the Medical Ethical Committee of Beijing Hospital of Traditional Chinese Medicine (2023BL02-054-01). This study is a retrospective investigation, the Ethics Committee has granted an exemption from the requirement for consent. In our research, data collectors have the capability to access potentially identifiable information about individual participants, such as their names. This access occurs during the data collection phase. We place significant emphasis on ethical considerations and employ anonymization techniques during data processing and analysis to ensure that individual identities are neither disclosed nor identifiable. Our study adheres to the principles of GCP and has obtained approval from the Research Ethics Committee of Beijing Traditional Chinese Medicine Hospital affiliated to Capital Medical University. We are committed to safeguarding the privacy of participants throughout the study, and our research report will not present any information that could reveal personal identities.

## Results

### Biological data

A total of 62 CNGM women were included in the study (Table 1). The mean age of these patients was 33.30 (range 24-54). A patient in her 50s had schizophrenia and had been taking olanzapine for a long time, and all the other patients were premenopausal women. The majority had a history of childbearing (98.39%). In 52 patients with detailed documentation of onset from the final delivery, all patients suffered the disease within eight years after the latest delivery, which 92.31% of patients were within six years after the latest delivery. There were 88.89% patients had a history of lactation, and the duration ranged from one to 48 months. Lactation disorders occurred on the unaffected side (19.23%) and the affected side (30.78%), respectively. There were 19.35% patients had GLM history before, 17.74% patients had a breast trauma history just before the onset of this disease, and 25.81% patients had hyperprolactinemia, including 1.61% patients had a pituitary tumors. There were 1.61% patients had psychiatric medication histories, and 3.23% patients had a family history of breast cancer.

**Table 1.**
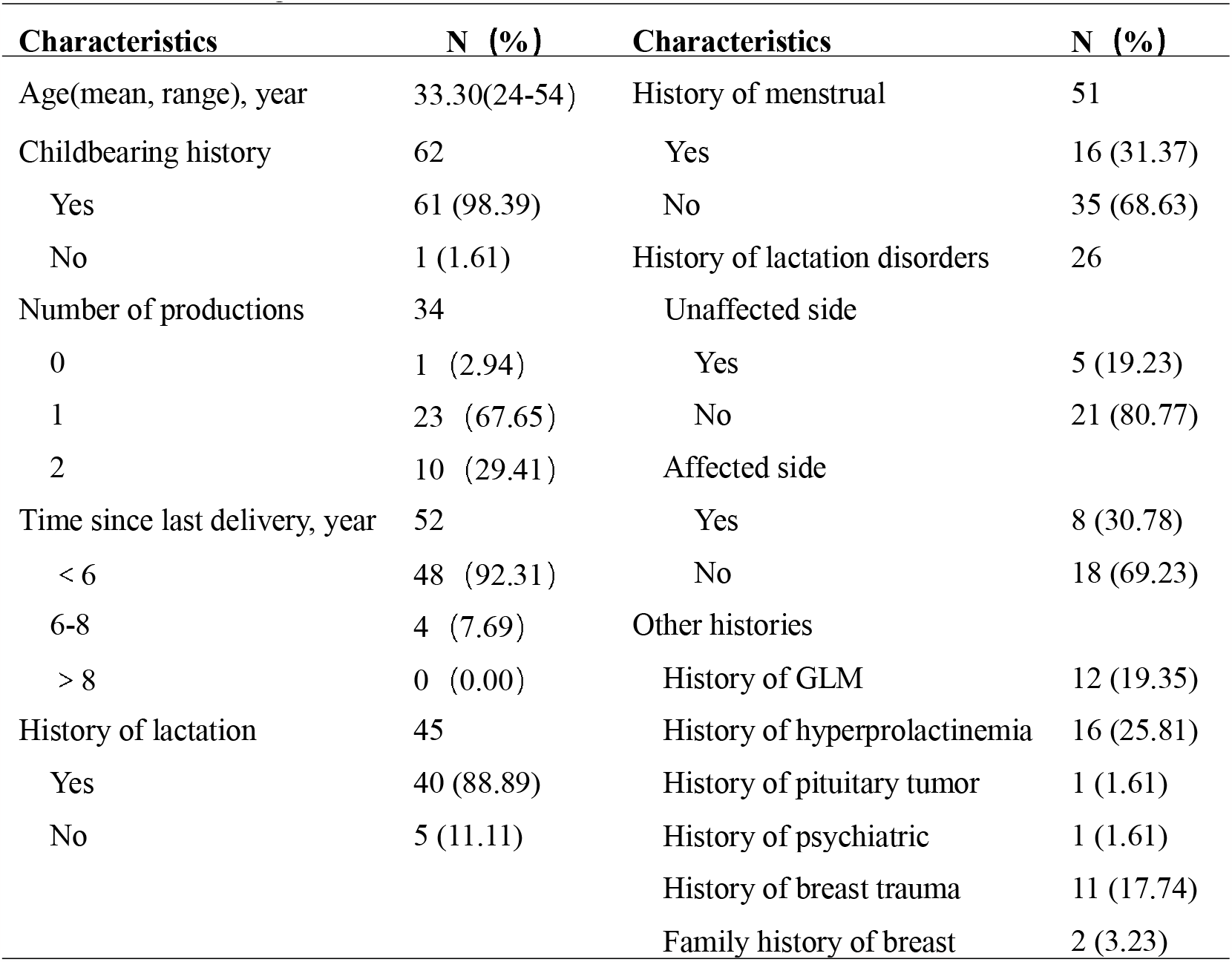
Biological Data of 62 Patients with CNGM.

### Clinical features

Clinical features are summarized in Table 2. Only 4.84% of patients were bilateral, and the others were unilateral. Pain and mass were the main manifestations of CNGM, and all patients (100.00%) had these two symptoms. The next symptoms were redness (72.58%) and ulcer (69.36%). Nipple retraction and discharge were seen in 33.87% and 14.52% of patients, respectively. Besides local signs, systemic symptoms were seen in some patients, including fever (8.06%), headache (1.01%), erythema nodosum (6.45%), and cough (4.84%). We performed a logistic regression analysis of nipple retraction and found it was an independent risk factor of CNGM. The OR value was 5.005 (Table 3).

**Table 2.**
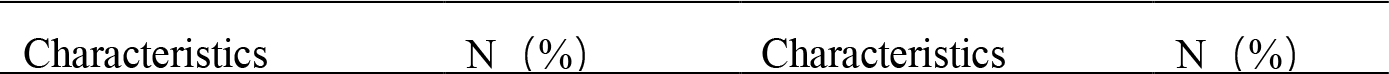
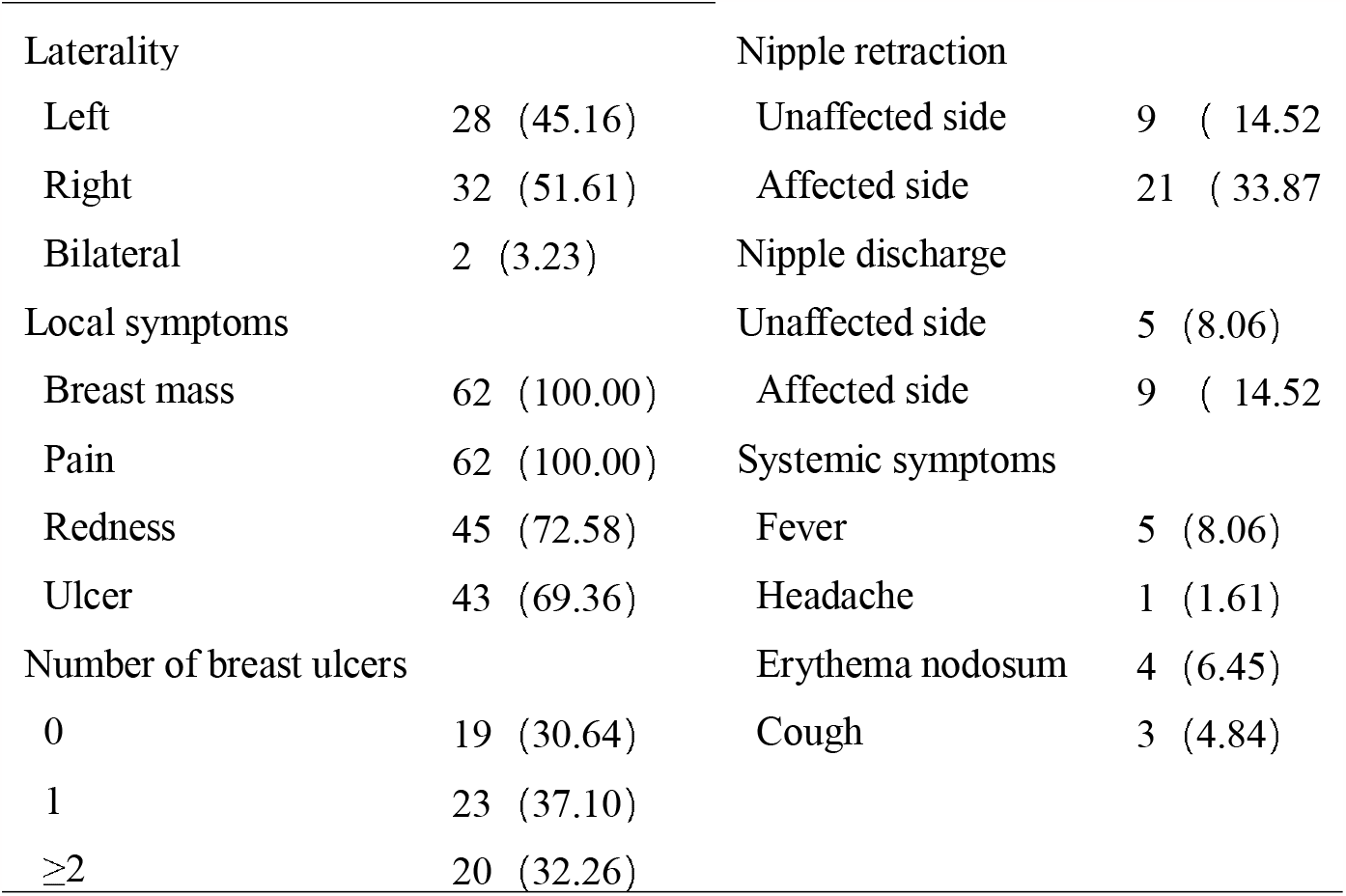
Clinical Characteristics of 62 Patients with CNGM.

|  |  |  |  |
| --- | --- | --- | --- |
| Laterality |  | Nipple retraction |  |
| Left | 28 (45.16) | Unaffected side | 9 (14.52) |
| Right | 32 (51.61) | Affected side | 21 (33.87) |
| Bilateral | 2 (3.23) | Nipple discharge |  |
| Local symptoms |  | Unaffected side | 5 (8.06) |
| Breast mass | 62 (100.00) | Affected side | 9 (14.52) |
| Pain | 62 (100.00) | Systemic symptoms |  |
| Redness | 45 (72.58) | Fever | 5 (8.06) |
| Ulcer | 43 (69.36) | Headache | 1 (1.61) |
| Number of breast ulcers |  | Erythema nodosum | 4 (6.45) |
| 0 | 19 (30.64) | Cough | 3 (4.84) |
| 1 | 23 (37.10) |  |  |
| ≥2 | 20 (32.26) |  |  |

**Table 3.** Nipple Retraction on the Pathogenesis of CNGM.

| Variable | Yes/No | Affected | Unaffected | OR | 95%CI |
| --- | --- | --- | --- | --- | --- |
| Nipple retraction | Yes | 22 | 9 | 5.005 | 1.966, |
|  | No | 21 | 34 |  |  |

aOR value > 1 indicates that the factor is a risk factor;

### Ultrasound

Hypoechoic areas could be seen in all 62 patients under ultrasound (100.00%), most of which were heterogeneous echo (hypoechoic with fluid zones) (56.45%). Nearly half of the patients had multiple hypoechoic areas (48.39%), and the hypoechoic areas were mostly subcutaneous (88.46%). Some patients were accompanied by axillary lymphadenopathy (59.68%), dilated ducts (47.39%), and thickening of the breast tissue (35.00%). (Figure 2 and Table 4)

**Table 4.** Ultrasound of 62 Patients with CNGM.

| Characteristics | N (%) | Characteristics | N (%) |
| --- | --- | --- | --- |
| Type of echoes | 62 | Subcutaneous involvement | 52 |
| Homogeneous echo | 27 (43.55) | Yes | 46 (88.46) |
| Heterogeneous echo | 35 (56.45) | No | 6 (11.54) |
| Number of echoes | 62 | Dilated ducts | 38 |
| 1 | 32 (51.61) | Yes | 18 (47.39) |
| ≥2 | 30 (48.39) | No | 20 (52.63) |
| Axillary | 62 | Thickening of the breast | 20 |
| Yes | 37 (59.68) | Yes | 7 (35.00) |
| No | 25 (40.32) | No | 13 (65.00) |

**Fig. 2.**
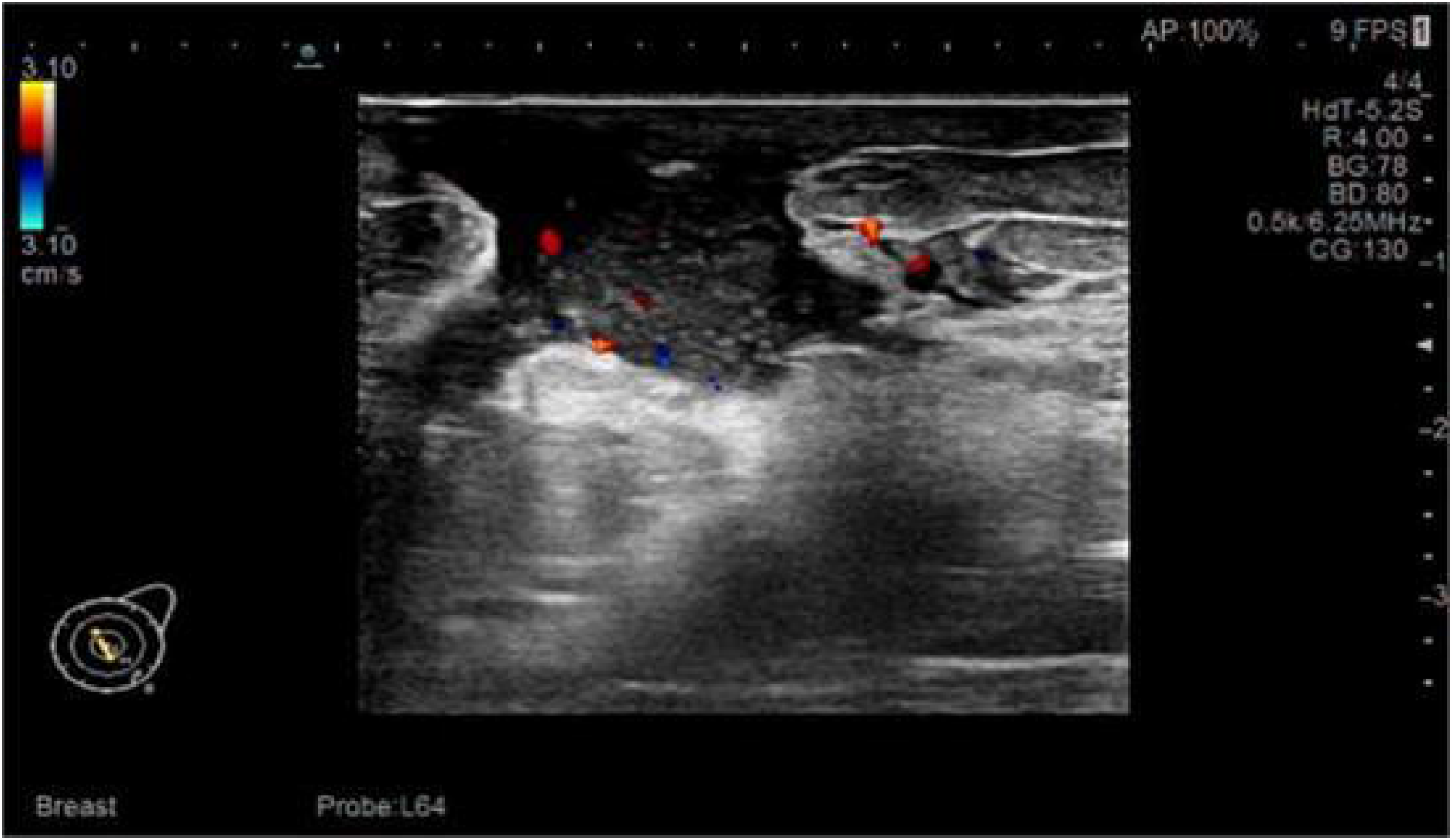
Manifestations of CNGM under ultrasound. (A)Irregular hypoechoic areas are on the medial side of the right nipple, with an an unclear border, no fluid dark zone is in it, and the skin is involved. CDFI: strip-shaped blood flow signals are in the center and perimeter. (B)The suitable breast skin is edema and thickened, and heterogeneous echoes are formed by multiple flaky hypoechoicic with a small amount of fluid dark zone, the skin is involved. CDFI: rich blood flow signals can be seen around and inside. (C)Enlarged eccentric target annular lymph node. (D)The milk duct is dilated to about 0.38 cm.

### Histopathology

Most of the pathologies of 62 patients were lobular structural destruction, acute and chronic inflammatory cell infiltration, and multinucleated giant cell reaction, accompanied by granulomas and cyst formation (Figure 3). The detection rate of gram-positive bacteria and gram-negative bacteria was38.71% and 8.06%, separately. Only two patients (3.23%) were detected, both gram-positive and gram-negative. (Table 5)

**Table 5.** Gram Staining of 62 Patients with CNGM.

| **Gram staining** | **N (%)** |
| --- | --- |
| Gram staining positive | 31 (50.00) |
| Gram-positive bacteria | 24 (38.71) |
| Gram-negative bacteria | 5 (8.06) |
| Gram-positive bacteria + gram-negative | 2 (3.23) |
| Gram staining negative | 31 (50.00) |

**Fig. 3.**
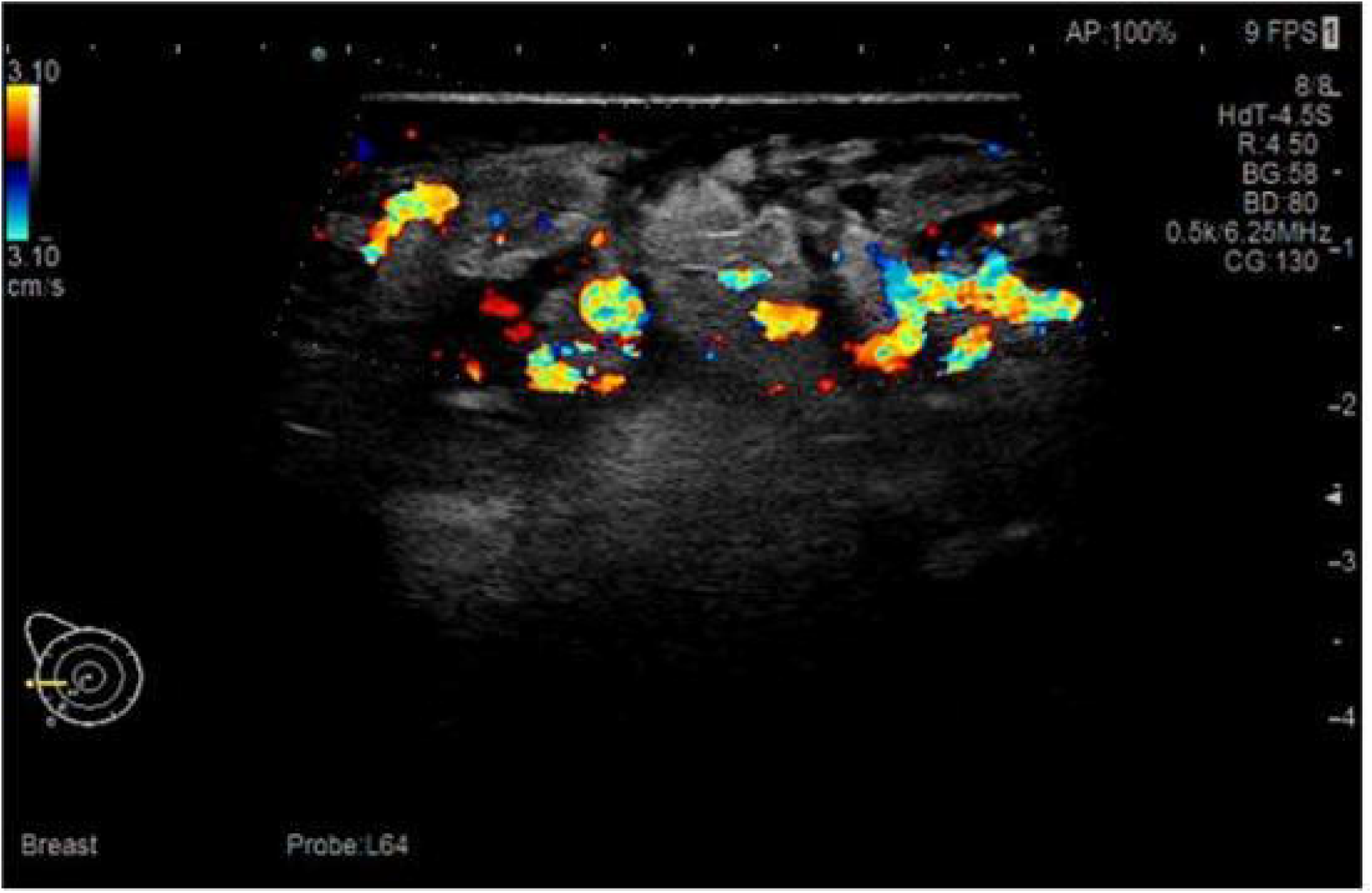
Morphological features of CNGM. (Gram staining: A×10 magnification, B×40 magnification),(Hematoxylin-Eosin staining: C×10 magnification, D×40 magnification). Lobular structure destruction can be seen in breast tissue, many acute and chronic inflammatory cells infiltrate, an interstitial granulomatous structure formed with multinucleated giant cell reaction, and mmicro abscesses and lipid vacuoles in some places(A, C). A single lipid vacuole surrounded by neutrophils and an outer cuff of epithelioid histiocytes, mixed inflammatory infiltrates including Langerhans giant cells and lymphocytes outside around it. Some bacteria can be seen in lipid vacuole (B, D).

## Discussion

Due to the lack of understanding of CNGM and the fact that previous studies are primarily small sample studies, the pathogenesis and clinical diagnosis of CNGM are still unclear. Very little data about its biological features, clinical characteristics, imaging manifestations, and histopathology were reported, so there is an excellent possibility of missed diagnosis and misdiagnosis of CNGM. A better understanding of CNGM may help improve the diagnosis and treatment, especially using antibiotics and immunosuppressants. This is the most extensive sample size study on the analysis of CNGM clinical features. The discussion will mainly focus on the biological data, clinical features, ultrasound manifestations, and histopathological features of CNGM.

### Biological data

CNGM, a benign breast disease with a unique histological structure, occurs mostly in women of childbearing age. Our result showed the average age was 33.3, consistent with the previous report that the onset ages range from 20 to 40^[1, 11]^.This study found that CNGM often occurred in women with a delivery history (98.39%). In 52 patients with detailed documentation of onset from the last delivery, all patients suffered the disease within eight years after the last delivery, and 92.31% of patients were within six years after the last delivery. Women with a breastfeeding history have the basis of milk stasis, especially within six years after delivery. Because of milk residue, the lobules and ducts may dilate and expand. So there are great possibilities that secretions cause local inflammatory response or skin inherent bacteria enter the breast lobule through the dilated duct to cause bacterial infection^[12-14]^. It is believed that milk residue and the occurrence of CNGM are correlated closely. Another evidence is that the lactation disorder that happened on the affected side (30.78%) is much higher than that on the unaffected side (19.23%). Poor milk discharge may increase milk accumulation, so it increases the chance of infection^[15]^.

Our study found that 25.81% of patients had hyperprolactinemia. The rate is much higher than in ordinary people. 1.61% of patients had pituitary tumors, and 1.61% had psychiatric drug intake history. We think it is reasonable to explain this phenomenon regarding both high prolactin levels and excessive secretion in the milk duct. High prolactin level has been shown to increase the risk of breast infections associated with inflammation response^[18-20]^, the patient with pituitary tumor had hyperprolactinemia, increased duct secretion, and eventually led the damage of the breast ducts. As we mentioned in milk residue, the reason is consistent with previous research reports^[3, 9, 18]^.

There was only one patient who did not give childbirth. Still, she had taken oral contraceptives for 2 months just before the onset of CNGM. Another patient in her 50s had schizophrenia and had been taking olanzapine long before the beginning of CNGM. Our findings showed psychiatric drug intake could increase the incidence of CNGM, too, because some antipsychotic drugs had the side effect of elevating prolactin levels or stimulating duct secretion. CNGM has been reported to have a higher incidence of psychiatric disorders in previous literature too^[9, 17]^. Diesing D et al. reported that some contraceptive components could cause endocrine disorders, which led to increased acinar secretions from the mammary glands, causing ductal endosecretion retention, duct dilation, which then duct rupture leading to persistent inflammation of stromal cells^[16]^. This can explain the reason why external hormone stimulation can result in CNGM.

In this study, 12 patients had granulomatous mastitis (19.35%) before; 7 cases occurred on the ipsilateral side and 5 cases on the opposite side, all of which recurred mastitis within two years. The higher occurrence has also been reported in other studies^[3, 4,21]^. The reason CNGM is accessible to relapse has not been noted before, and we tried to explain it from 3 aspects. (1) Anatomy factors: These patients may have unique breast structure characteristics, such as nipple retraction and slender and twisted milk ducts. This is likely to cause duct obstruction and breast secretions accumulation, resulting in mammary infection. Nipple retraction is a risk factor, as we have proved before. Previous studies have also confirmed that some non-puerperal mastitis (NPM) occurrences and recurrence are related to breast duct abnormality and secretion accumulation^[22]^. (2) Hormone factor: Yajing Huang et al. ^[23]^ emphasized that the disorder of prolactin level is an independent prognostic factor for the recurrence of breast inflammation High prolactin level was found in 25.8% of patients with CNGM in our study which was much higher than in average population. (3) Bacterial colonization factor: bacterial colonization is also an essential factor in the recurrence of mastitis. A small number of bacteria may increase to large quantities under the suitable conditions created by milk stasis and cause the relapse of inflammation. As a part of the normal skin microbiota, corynebacteria’s pathogenicity has been overlooked. As a corynebacterial, Corynebacterium kroppenstedtii differs from most other corynebacteria in that it requires a lipophilic environment to flourish. In recent years, more and more studies have shown that a good lipid environment of milk provides the basis for C. kroppenstedtii’s growth. There is a specific relationship between C. kroppenstedtii and mastitis^[1, 19, 24]^.

There were 17.74% patients had a history of breast trauma, in which 14.52 % developed CNGM, and 3.23% showed worsening clinical symptoms after the trauma. Based on the considerable tension induced by milk stasis and breast duct dilation, the appearance of trauma can be the last straw to cause duct rupture, the secretions in the duct will subsequently overflow to the mesenchyme, resulting in local acute inflammatory reaction. Previous studies have identified breast trauma as one of the risk factors for lactational mastitis, too^[25, 26]^. Our research shows the same result in CNGM, the specific NPM.

Compared with the crude global incidence of breast cancer, 58.5/100000^[27]^, 3.23% of patients had a family history of breast cancer with CNGM is much higher. As an inflammation-related disease, mastitis has rarely been considered a predisposing factor for breast cancer^[28]^. Paradoxically, previous epidemiological studies have established chronic inflammation related to many cancers, such as stomach, esophageal, and colon cancers^[29-31]^. Studies by Peters F, Lambe M, Ghadiri F, Chang C-M, et al. elucidated a possible association between mastitis and breast cancer, which all suggested an increased incidence of breast cancer in women diagnosed with chronic mastitis^[32-35]^.

The relationship between breast cancer family history and mastitis is rarely described in the existing literature. Based on the above reports, we try to understand their relationship from three aspects. First, mastitis and breast cancer are related to cytokines including IL-6, IL-8, TNF-α, NF-ĸB, etc. The transcription and expression of these cytokines form a mutual feedback loop by promoting cell proliferation and necrosis, generating an environment that promotes inflammation and tumors together^[36]^. Second, there are exogenous and endogenous pathogen-associated molecular patterns (PAMPs) (bacteria, viruses, and fungi, as well as endogenous molecules released by damaged or dead cells) in the activation of the CNGM inflammatory pathway, long-term exposure to these PAMPs may damage breast tissue and increase the risk of breast cancer^[37]^. Third, it is confirmed that some microbe infections related to chronic inflammation could reduce cancer, such as Helicobacter pylori (gastric adenocarcinoma) and enteric salmonella serotype Typhimurium or paratyphoid (cholangiocarcinoma) ^[36, 38, 39]^. It provides an idea for us to understand the correlation between C.k related CNGM and breast cancer. In addition, we also consider understanding this phenomenon at the genetic level: breast cancer and mastitis may have overlapping pathogenic genes, but these genes can modulate the pathogenesis of CNGM and breast cancer together. Whether CNGM would promote tumorigenesis or has a higher incidence of breast cancer can be further explored.

Sixteen patients had a menstrual irregularity history (31.37%). Menstrual irregularity often indicates endocrine dysfunction, including hormonal disorders. Breast tissue has been affected by endocrine hormones^[40]^. There is no systematic study on the relationship between endocrine disorders and NPM. Our future study will include research on the potential connection between CNGM and endocrine disorders.

Based on the above biological data, high-risk factors of CNGM includes child delivery history, especially women 6 years after delivery, lactation disorder, high prolactin level, breast trauma, and breast cancer family history. The first four correlate to duct secretion accumulation and mammary duct dilation, which can cause an inflammatory response. External trauma is another predisposing factor that can cause breast duct damage, even rupture, especially when there are significant tensions in the mammary duct. Anatomical and individual genetic differences may play a role in pathogenesis.

### Clinical features

Among the 62 patients in this study, 95.16% were unilateral, and three patients (4.84%) were bilateral. We found that the two patients with bilateral breasts were CNGM, and another patient was diagnosed with CNGM in the right breast and plasma cell mastitis (PCM) on the left side. This kind of case has not been reported before.

This study showed all patients with CNGM had different degrees of pain and breast mass (100.00%). Breast cancer is generally manifested as a painless mass. We believe that the pain of breast masses can be used to distinguish CNGM from breast cancer, although they are highly similar in some clinical and imaging features. Their definitive diagnosis and differentiation still need histopathological results.

There were 72.58% patients had breast redness. This is a performance of acute inflammatory reaction of CNGM. A total of 43 patients presented with ulcers (69.36%), of which 20 patients had multiple ulcers (46.51%). Most of the previous reports described breast ulceration, but the ulcer number was not counted. As we know, this study is the first to report breast ulcer numbers of CNGM. The ulcer number in CNGM is significantly increased compared with that in other NPM. Prior studies have shown that patients with bacteria infection were more likely to have breast ulceration and sinus tracts^[41]^, this explains why CNGM has more ulcers. (bacteria data are shown in 3.4)

14.52% of patients had nipple discharge on the affected side, suggesting that in addition to the common breast masses, pain, and redness, nipple discharge is another important clinical manifestation of CNGM. It is believed to be related to increased prolactin levels. The proportion of nipple retraction on the affected side (33.87%) and the unaffected (14.52%) side was quite different, and the results of logistic regression suggested that nipple retraction was a risk factor for the occurrence of CNGM, nipple retraction may cause poor milk discharge and secretions retention. It is worth noting that nipple retraction in CNGM patients is mainly congenital, which differs from that in breast cancer.

Some patients were accompanied by constitutional symptoms such as fever (8.06%), headache (1.01%), erythema nodosum (6.45%) and cough (4.84%). Headache occurred in 1 of 5 febrile patients, which may be one of the systemic inflammatory reactions caused by local inflammatory stimuli.

Previous investigations of CNGM have also mentioned the feature of erythema nodosum^[42]^. Given that erythema nodosum is mostly secondary to various autoimmune diseases^[43]^, erythema nodosum and cough may be the systemic signs that indicate CNGM is an autoimmune disease. Studies have reported that patients with CNGM have achieved better therapeutic effects after taking steroids or immunosuppressive drugs such as methotrexate, confirming that CNGM is an autoimmune disease^[44, 45]^.

The clinical characteristics are concluded: Breast masses and pain are the most common symptoms in all CNGM patients, followed by local redness and ulcer of the breast, nipple retraction, and nipple discharge can be seen in some patients. Systemic symptoms, including fever, headache, erythema of the limbs, and cough, are common in CNGM patients.

### Ultrasound

Ultrasound was the preferred imaging modality in breast examination. Ultrasonic characteristics of CNGM were rarely reported in the literature. The most common findings were mass, dilated ducts, edema, abscess, and effusion, with BI-RADS scores ranging from 2 to 5^[1, 9, 21]^. Breast ultrasound for all patients in this study showed hypoechoic areas; 48.39% showed mixed echo areas.

In contrast, breast cancer often presents as a single hypoechoic mass, which also helps us differential diagnosis. There were 56.45% patients presented as hypoechoic areas with fluid zones, and the probability of sinus and fistula in the later stage was higher. There were 88.46% of patients had subcutaneous involvement, which was redness and swelling in the skin. There were 68% of patients had axillary lymphadenopathy, which we believe is a breast inflammation response.

### Histopathology

Most of the pathologies in this study are lobular structural destruction, acute and chronic inflammatory cell infiltration, and multinucleated giant cell reaction, accompanied by granulomas and cyst formation, a central lipid vacuole surrounded by neutrophils and an outer cuff of epithelioid histiocytes can also be seen. Many scholars believe that CNGM is closely related to Corynebacteria. The detection rate of Corynebacterium varies from 10.5% to 100%, some of which were found in retrospective studies^[3-6, 17, 21, 44, 46-48]^. The most common isolates were Corynebacterium kroppenstedtii^[2, 44]^. The presence of gram-positive bacteria was detected in 26 patients (41.94%) in our study, and the detection rate of gram-positive bacteria in former studies was 36.8%-100%^[3, 11]^. Tariq H et al. surveyed 67 patients with GLM as the case group and 10 with non granulomatous breast abscess as the control group. Only 17.9% of 67 GLM cases detected the presence of gram-positive bacteria, and subsequent studies have demonstrated that all 12 gram-positive cases belonged to CNGM. All 10 patients in the control group had negative Gram stains and did not show features of CNGM^[49]^. Combined with the literature^[50]^, it can be seen that CNGM and GLM have significant differences in the detection rate of gram-positive bacteria. Many scholars believe that the detection rate of related pathogens such as Corynebacterium and other gram-positive bacteria is far lower than it is. We have combined previous research to summarize the reasons: First, pathogens in CNGM, including Corynebacterium, are picky bacteria with high requirements for culture conditions and culture time^[1]^. The detection rate of pathogens is affected by the methods used. The detection rate of gram-positive bacteria increased by 21% by using 6 μm thick sections rather than the standard 4 μm one in Gram stain^[11]^. Second, compared with samples from the operation, this study’s sample size is limited because most of them were obtained by the core needle. Third, some patients may have been empirically treated with antimicrobial therapy before puncture, resulting in the underestimation of pathogens. Fourth, pathogens such as Corynebacterium, as members of the intrinsic flora on the skin surface, may be considered contaminants and neglected too. The above reasons make detecting CNGM pathogens more complex in clinical practice. The detection of gram-negative bacteria is rarely described in the existing literature. Gram negative bacteria was detected in seven patients. In addition, some studies have detected the presence of nontuberculosis mycobacteria, Staphylococcus aureus, coagulase-negative staphylococci, and Escherichia coli^[17, 51]^. Further research on suspected pathogenic bacteria is significant for clinical antibiotics and other treatments.

## Conclusions

The results of this study have unveiled several factors associated with CNGM. These factors include the presence of milk residue and excessive duct secretions, ex-ternal trauma, hormonal disorders such as hyperprolactinemia, and bacterial colonization. These elements appear to play a pivotal role in both the onset and recurrence of CNGM. Clinical manifestations commonly involve pain and the development of breast masses. These symptoms can also be accompanied by local indicators like redness, ulceration, nipple discharge, and nipple retraction. Furthermore, systemic symptoms, including fever, headache, erythema nodosum, and cough, may also manifest in some cases. Ultrasound findings in CNGM cases predominantly show hypoechoic masses and dilated ducts, along with thickening of the breast tissue, subcutaneous involvement, and axillary lymphadenopathy. Interestingly, the detection rate of gram-positive bacteria in CNGM cases stands at 41.94%, which is significantly higher compared to other non-puerperal mastitis (NPM) conditions, such as granulomatous lobular mastitis (GLM). Based on the identified predisposing factors, tailored treatment strategies can be employed. These strategies include interventions like the correction of inverted nipples, the administration of antibiotics, and the implementation of prolactin-reducing approaches, such as the use of Bromocriptine. This is particularly important for patients with elevated prolactin levels.

However, it is important to acknowledge certain limitations in this study. The relatively limited duration of data collection, as well as the exclusive use of data from our hospital, has resulted in an insufficient number of cases. Furthermore, the absence of control groups, such as GLM or healthy women, has contributed to the incompleteness of available information. As a retrospective study, there were some imprecisions in the data. Although menstrual disorder data were collected, hormonal parameters, including progesterone and estradiol levels, were not tested in this study. These hormone levels will be subject to analysis in subsequent research. To enhance the robustness of these findings, it is imperative to consider a larger sample size and include control groups in future investigations. The hope is that the results of this study will contribute to a deeper understanding of CNGM and pave the way for the development of more effective treatment strategies in clinical practice.

## Acknowledgments

All authors made a significant contribution to the work reported, whether that is in the conception, study design, execution, acquisition of data, analysis and interpretation, or in all these areas; took part in drafting, revising or critically reviewing the article; gave final approval of the version to be published; have agreed on the journal to which the article has been submitted; and agree to be accountable for all aspects of the work.

## Institutional Review Board Statement

This study was approved by the Medical Ethical Committee of Beijing Hospital of Traditional Chinese Medicine (2023BL02-054-01).

## Informed Consent Statement

Informed consent was obtained from all subjects involved in the study.

## Data Availability Statement

The original contributions presented in the study are included in the article. Further inquiries can be directed to the corresponding authors.

---

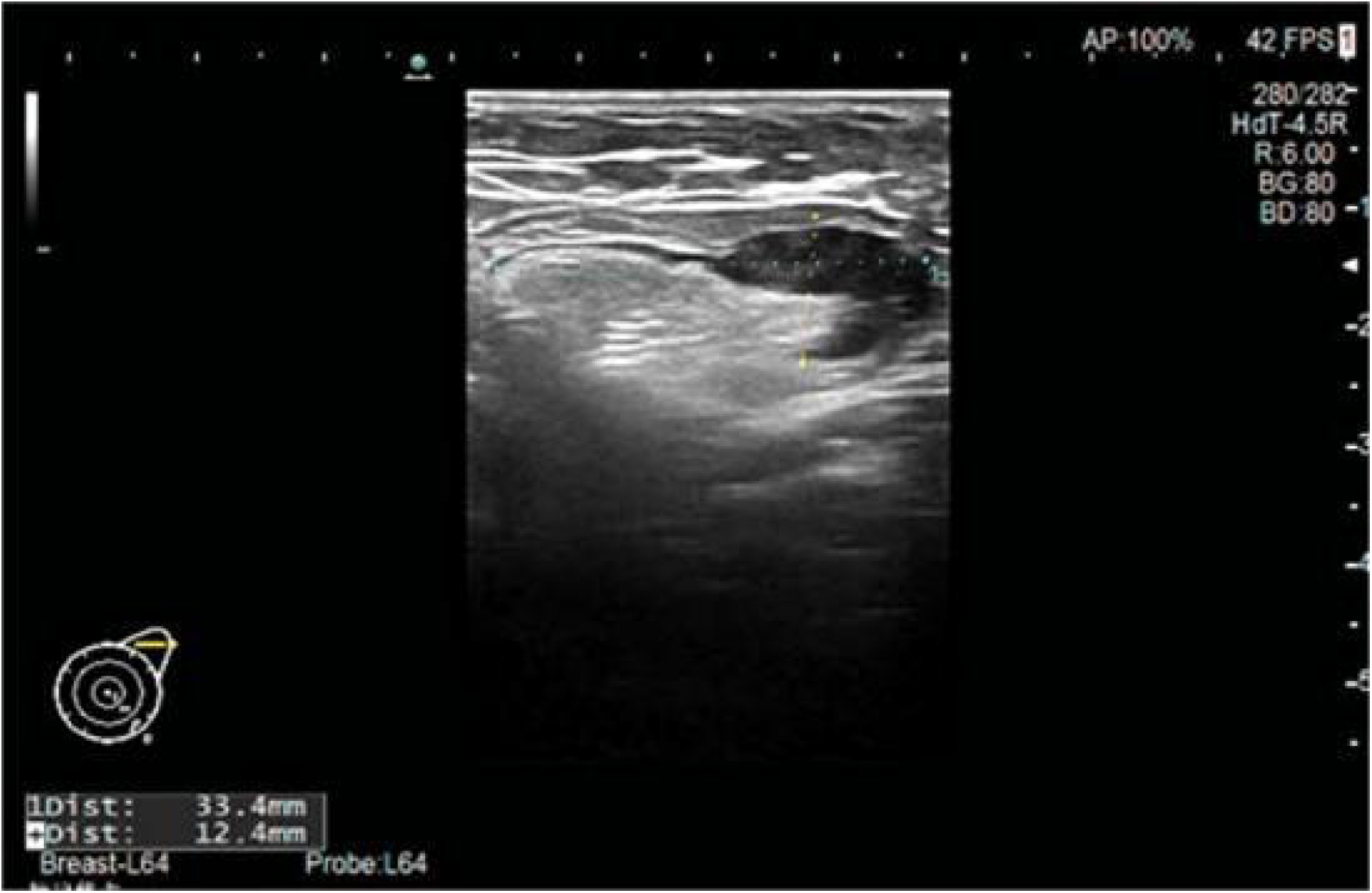

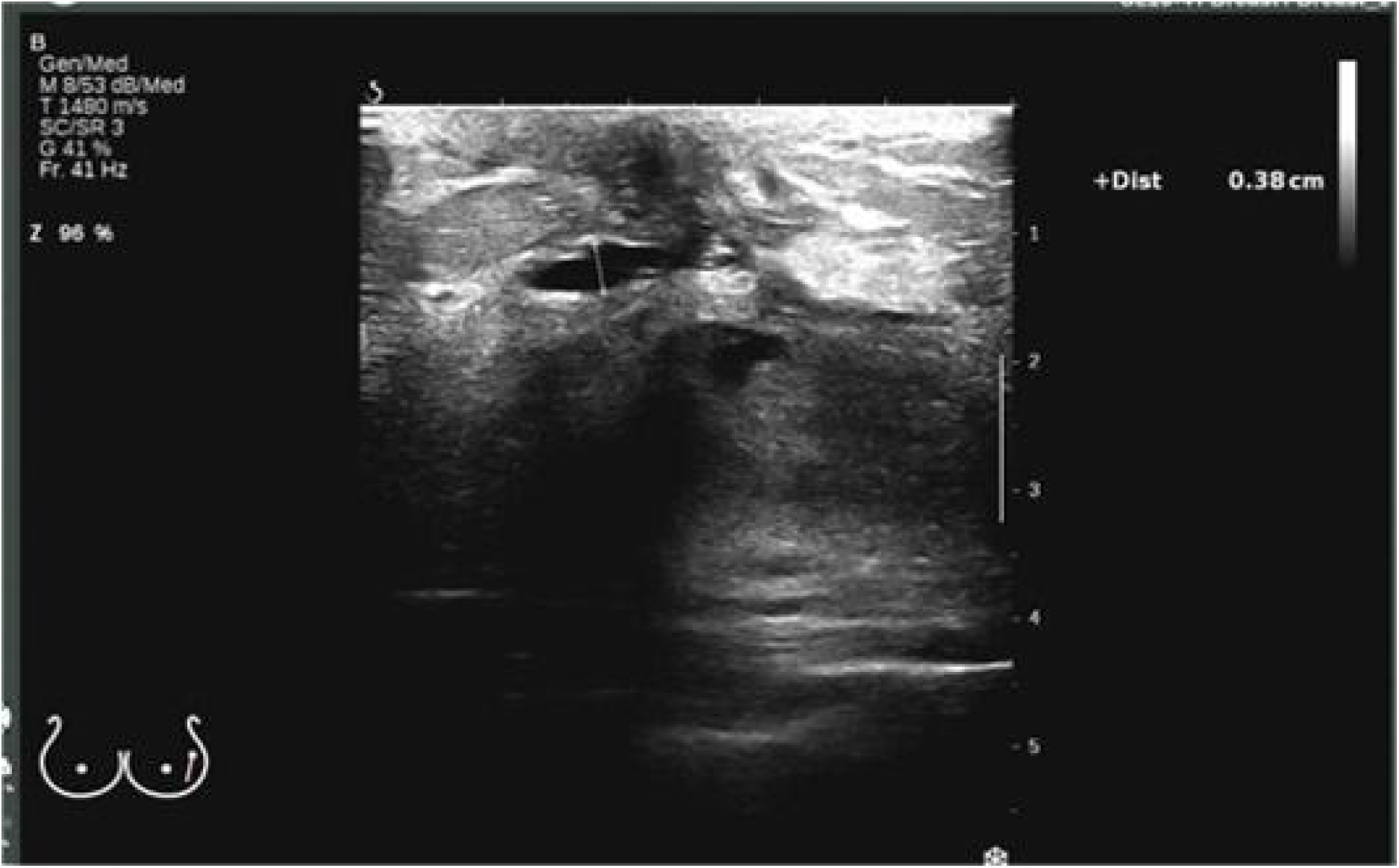

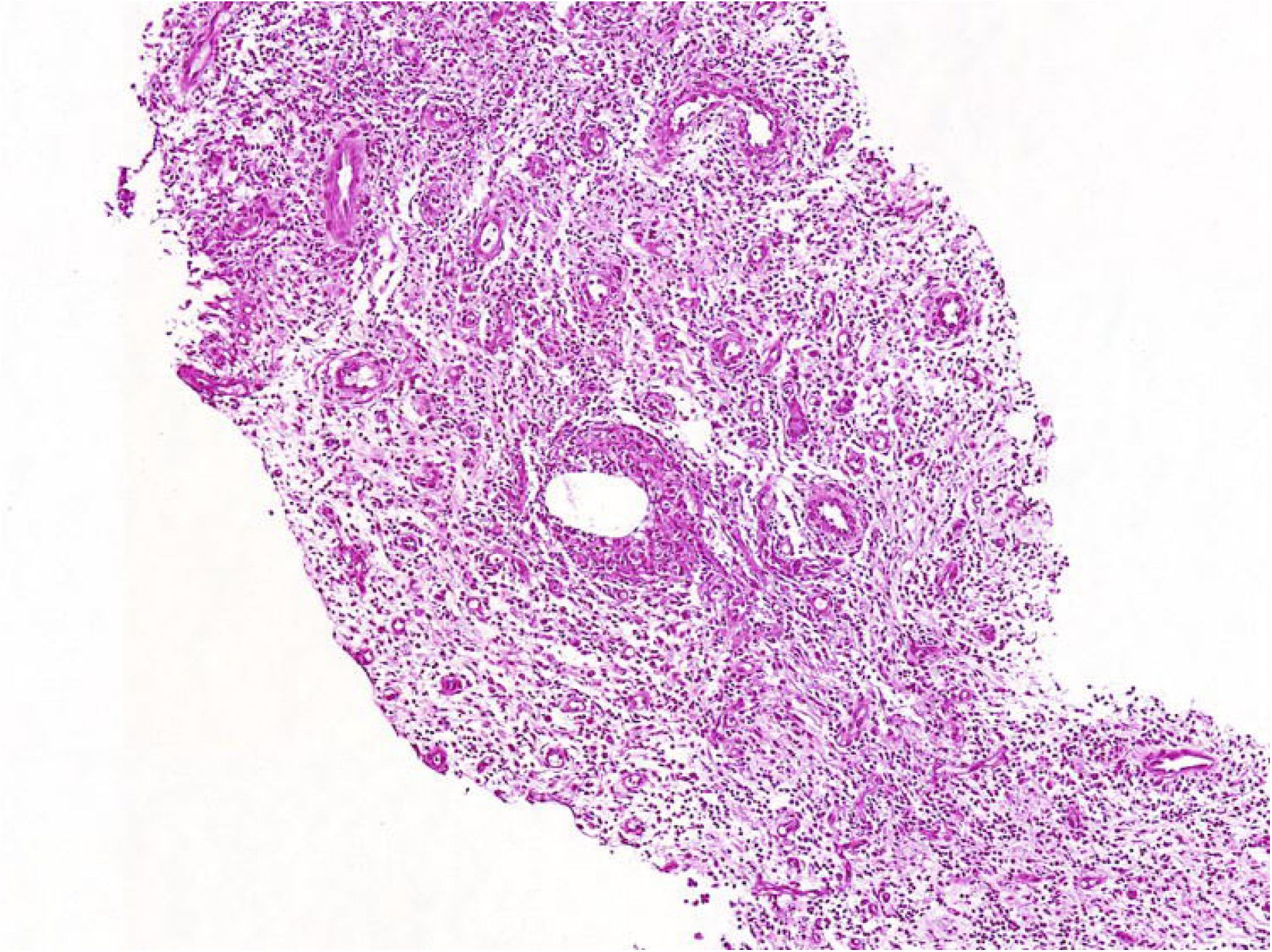

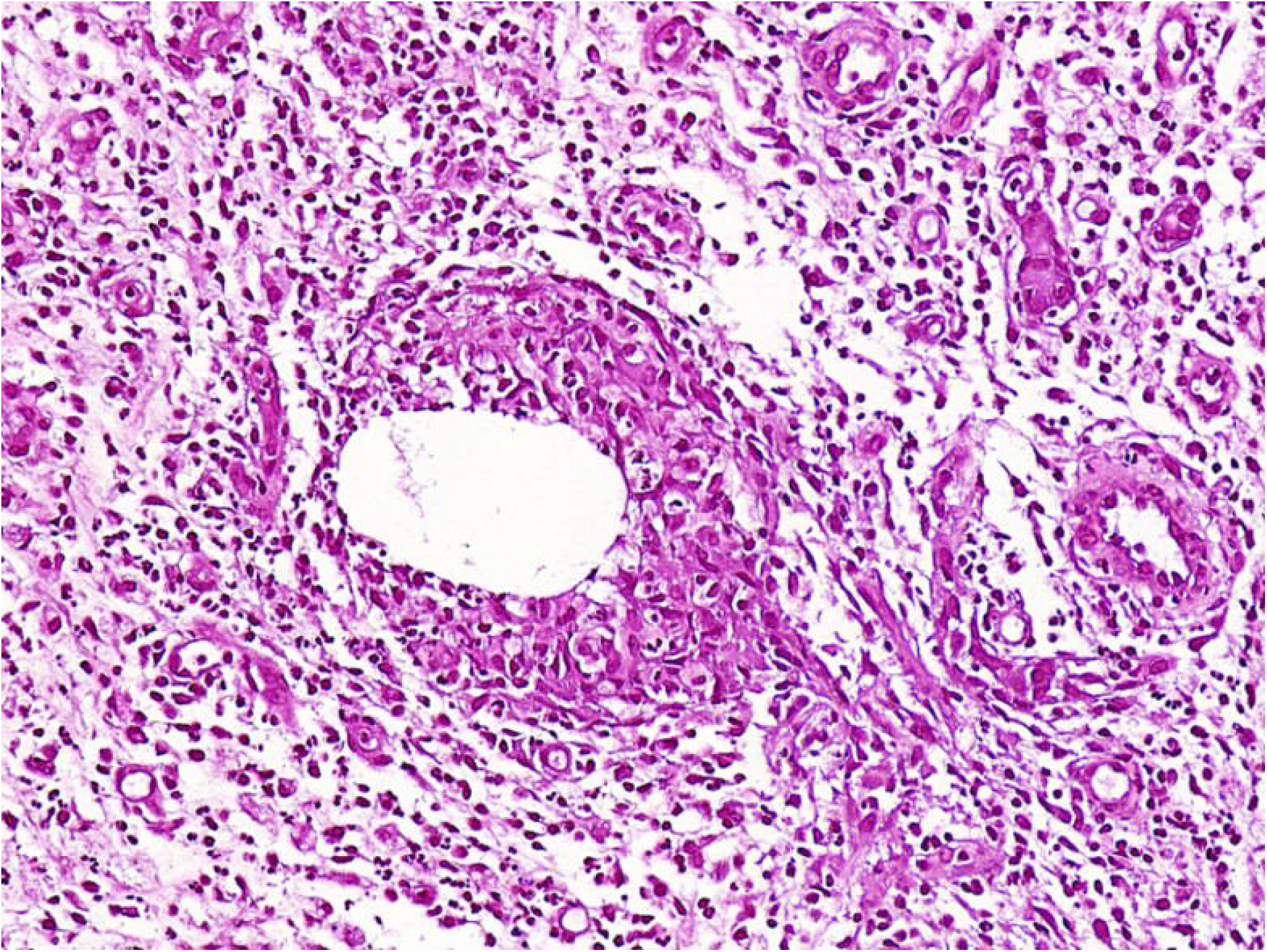

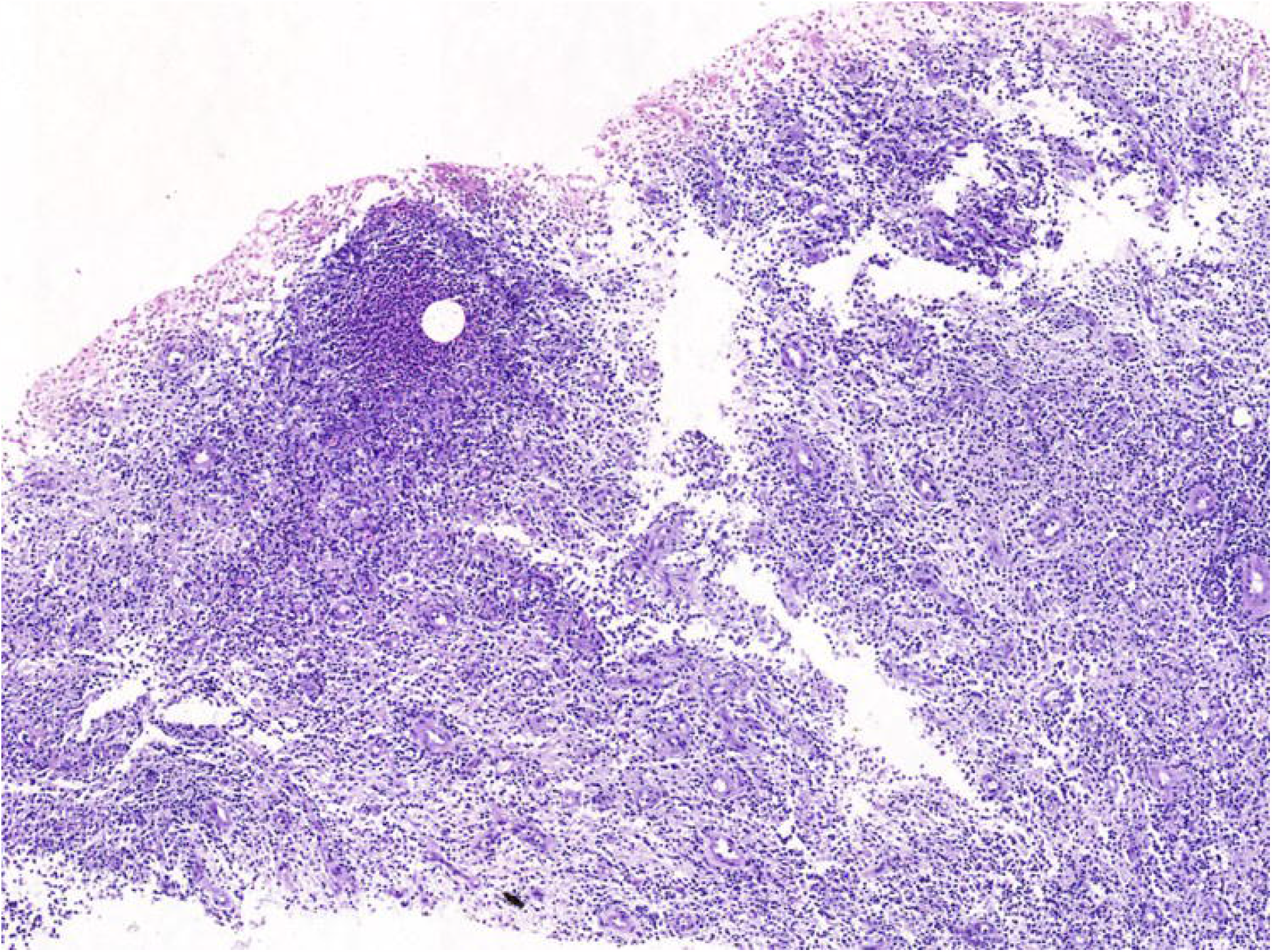

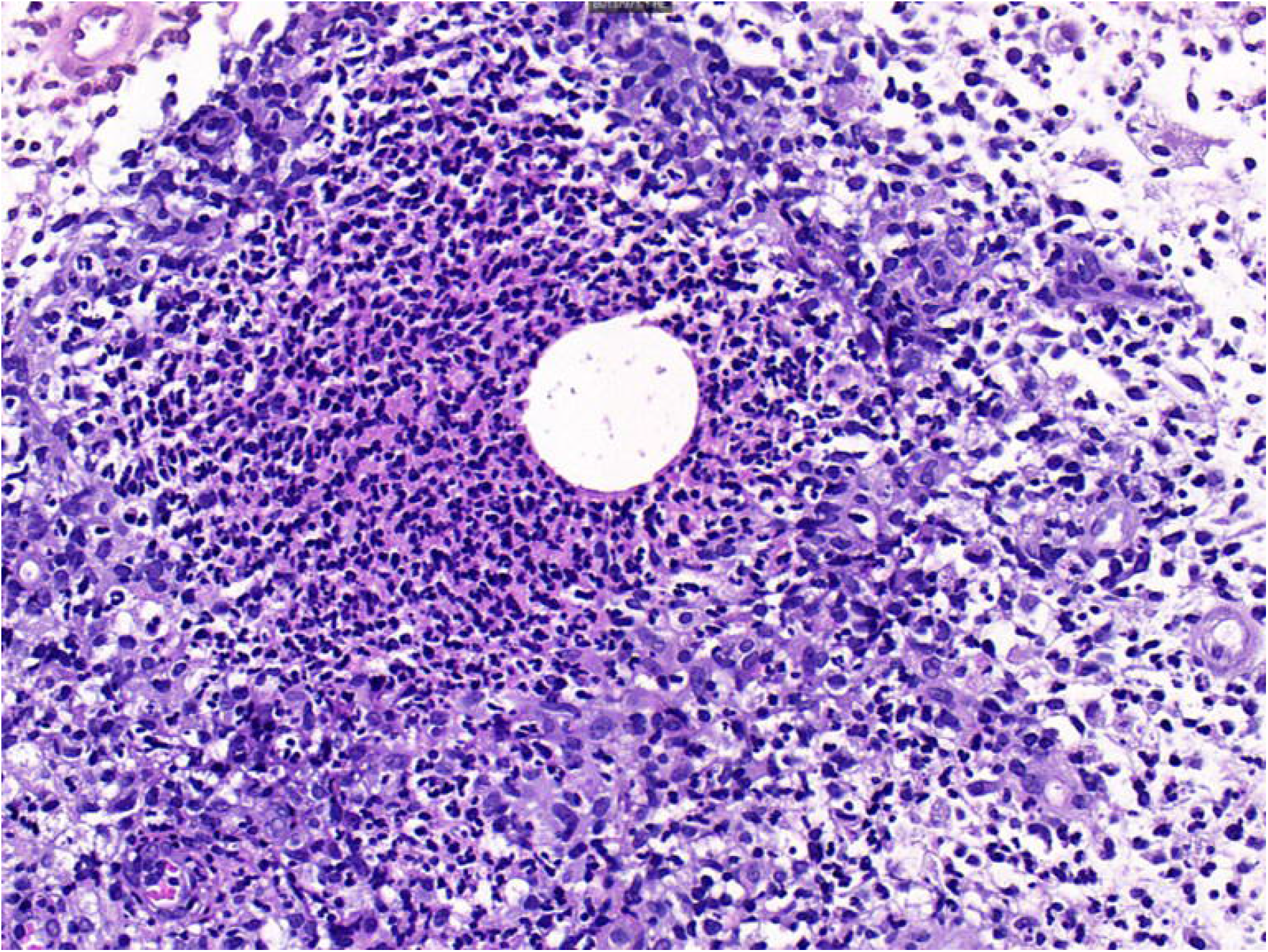

